# Readmission after Heart Failure Hospitalization: An Environmental Scan of Alberta Initiatives and Outcomes

**DOI:** 10.1101/2025.09.24.25336368

**Authors:** Camilia Thieba, Natalie Wiebe, Chantal Atwood, Zandria Morley, Alexis Guigue, Robin L Walker, Jonathan Howlett, Hude Quan, Cathy A. Eastwood

## Abstract

This environmental scan aims to identify and describe initiatives implemented in Alberta, one of the few Canadian provinces with a unified health delivery system, to reduce heart failure (HF) readmissions. It also acknowledges the challenges in attributing direct benefits to these interventions.

Using snowball sampling, we identified and recruited 11 employees and clinicians from Alberta Health Services (AHS) who possessed significant historical institutional knowledge about HF. Academic and grey literature were reviewed related to Alberta’s readmission reduction initiatives and reported outcomes. Unstructured, in-depth interviews were conducted to clarify timelines and provide detailed descriptions of these interventions.

Our findings indicate substantial clinician efforts over 15 years to address all-cause readmissions post HF hospitalization in Alberta, encompassing a range of interventions from small-scale projects to large multi-city, multi-stakeholder initiatives. Assessing the impact of smaller interventions on provincial readmission rates proved challenging; however, five major initiatives collectively led to a 1.8% reduction in 30-day all-cause readmissions province-wide (from 22.2% to 20.4$, p=0.04). Key factors that appeared to support these efforts included utilization of the EMR system, stakeholder engagement in standardized care, effective communication practices, and appropriate resource allocation. Clinical teams are now integrating successful components from these initiatives into the province-wide clinical information system, Connect Care, to enhance care coordination and patient outcomes.

This environmental scan highlights various comprehensive initiatives in Alberta aimed at improving patient care and reducing readmission after HF hospitalization. While an overall 1.8% reduction in readmission rates was observed over the 15 years, attributing this change directly to the interventions is challenging due to various implementation barriers and the complexity of healthcare delivery.

Continued efforts towards personalized care and innovative EMR utilization hold promise for further improvement in HF readmission rates.

## Introduction

Heart Failure (HF) is a chronic progressive condition with acute decompensation episodes. It involves structural and functional weakening of the heart, leading to reduced cardiac function and subsequent poor quality of life[1]. In 2022, the Canadian Institute for Health Information (CIHI) reported HF as the third leading cause of hospitalization across all Canadian populations[2]. Care for HF patients is not only costly but also challenging due to multiple comorbid conditions and associated frailty[3]. In Alberta, where 2% of adults live with HF and 1 of 5 patients are readmitted within 30 days of discharge, HF management costs the provincial healthcare system close to $100 million annually[4,5]. In addition to these costs, each readmission further deteriorates the quality of life for patients with an already frail disposition[6].

To address this problem, several jurisdictions initiated projects over the years to improve patient outcomes. In Canada, since 2009, the province of Alberta has established a unified zone-led health delivery system through Alberta Health Services (AHS) (Figure 1)[7]. To reduce readmission rates across the province, the main AHS strategy has been standardization and integration of care.

**Figure 1.**
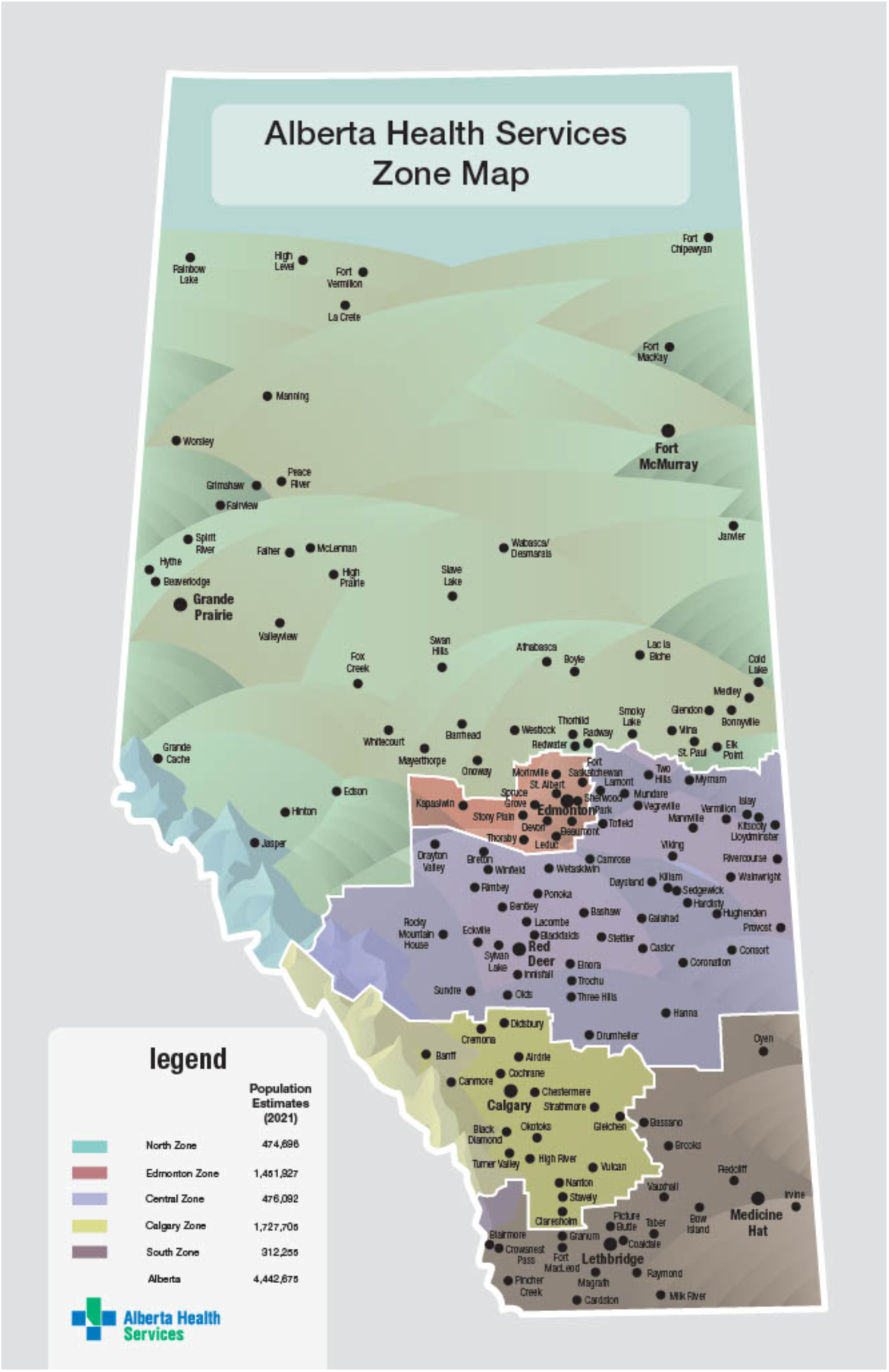
Alberta Health Services Zone Map (Alberta Health Services granted permission to reproduce this figure on January 17, 2022). Alberta Health Services. Alberta Health Services Zone Map. Available at: https://www.albertahealthservices.ca/assets/zone/ahs-map-ahs-zones.pdf. Accessed on July 25, 2022.

Since the results of these initiatives are not fully explored or publicly reported, their impact on patient outcomes remains largely unknown. In recent years, the province has embarked on a substantial health data integration initiative, Connect Care, whereby Epic (Epic Systems Corp., Verona, WI) clinical information system is being deployed province-wide, to address healthcare system fragmentation, evaluate care pathways, and centralize electronic health data. Therefore, we sought to gain a clearer understanding of past efforts to decrease HF readmission rates in Alberta, in hopes of applying key strategies to the newly designed Connect Care HF pathway. This pathway is meant to implement research findings into a series of physician order sets, dashboards, and clinical decision support tools, to optimize HF care across Alberta. Accordingly, the purpose of this paper is to 1) describe and explore the initiatives over time to reduce HF readmission rates since the amalgamation of AHS from discrete Health Regions, and 2) describe all-cause 30-day readmission rates within Alberta over the study period.

## Materials and Methods

### Literature Search Methods

To gain insights into the structures, processes, and outcomes of projects in Alberta, the team scanned the medical peer-reviewed literature between Jan 1, 2008, to Dec 2021. The topic keywords included “heart failure”; “disease-management-programme”; “hospital readmission”. Four databases were searched: PubMed, PsycInfo, CINAHL, and Google Scholar. The search was limited to publications about Alberta. To scan the grey literature, we gathered documents from the AHS website, Clinicaltrials.gov, and ProQuest Dissertations & Theses, as well as from co-authors, and associated experts. The databases and websites were searched as in the PRISMA diagram (Figure 2). We included papers published in English describing the efforts undertaken in Alberta to reduce readmissions after HF hospitalization.

**Figure 2.**
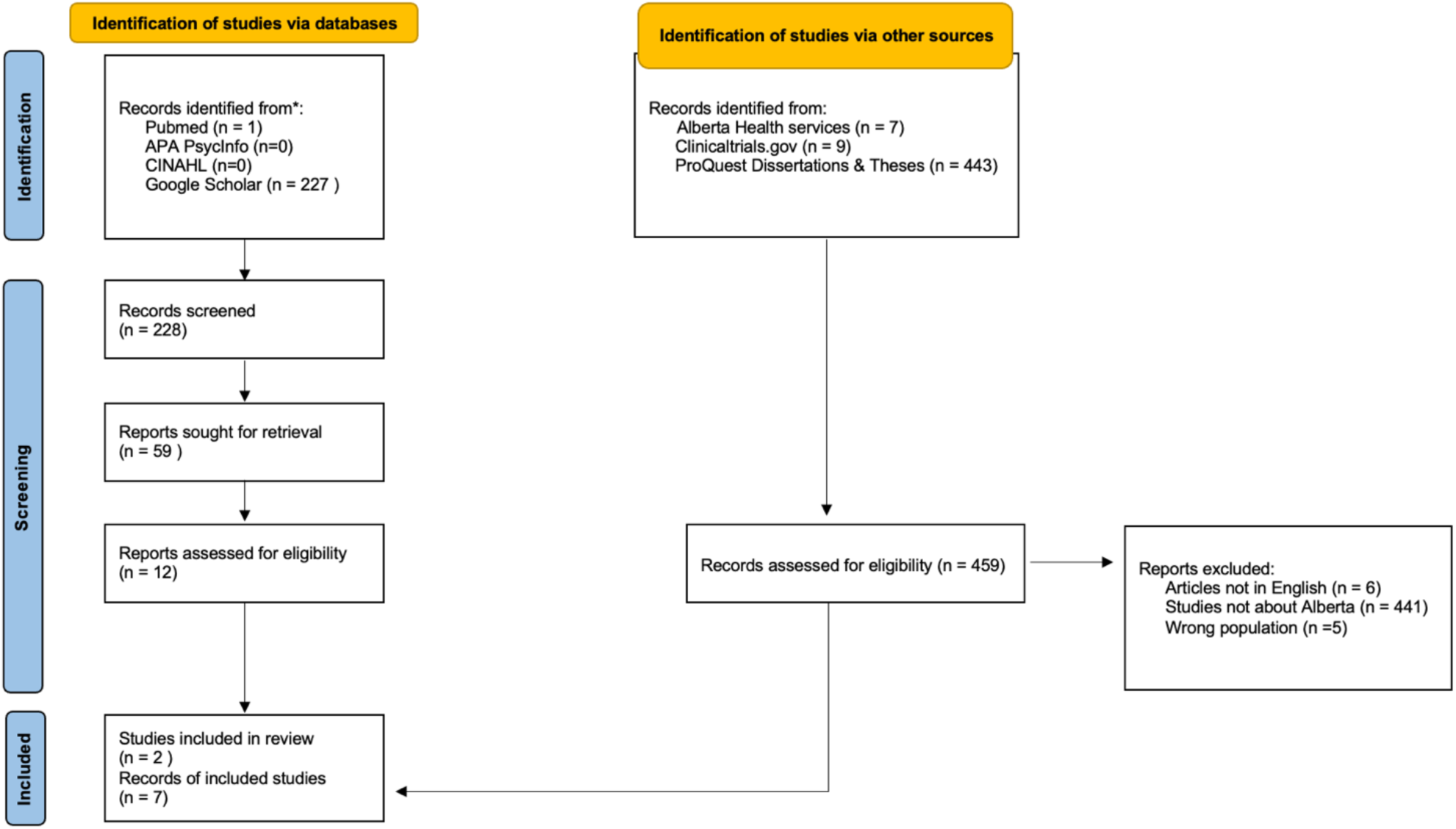
PRISMA Flow Diagram of Peer-reviewed and Grey Literature Searched in 2020.

### Qualitative Methods

We took an inventory of HF initiatives conducted in Alberta between 2009 and 2020. The scope of our analysis was restricted to the major projects for which information was available. Primary sources of information for this environmental scan were interviews with key informants (current or former employees of AHS) with extensive historical knowledge of the HF projects in Alberta.

Secondary sources of information included documentation provided by participants or co-authors in addition to the peer-reviewed and grey literature about HF readmission reduction initiatives in Alberta. The information gathered focused on structures (settings and policies), processes (interventions), timelines, and outcomes related to HF care in Alberta since 2008, based on a framework for medical quality of care assessment[8].

### Semi-Structured Interviews with Key Informants

Using purposive sampling[9], we identified and recruited key informants comprising one clinician and one analyst who were closely involved in foundational HF patient improvement projects. From these individuals, we employed snowball sampling to identify other physicians and AHS managers, including nurses, who had been closely involved in heart failure patient flow and readmission reduction initiatives over the past 15 years. All 11 participants were selected because they were heavily involved from the start and contributed to most, if not all, of the interventions. This approach ensured that those interviewed had direct and comprehensive insights into the development and implementation of these initiatives.

Semi-structured interviews were conducted between December 2019 and June 2020 by CE, AG, and CT. Informed consent was obtained prior to each interview. Ethics was obtained from the Conjoint Health Research Ethics Board REB20-0684. The 60-minute interviews involved videoconferencing and/or in-person meetings. After each interview, major initiatives were reviewed with the participants by email and further clarification was provided if needed. Interviews continued until a clear understanding of timelines, structures, and processes was reached. Qualitative description involves content analysis, a method for fact-finding and summarizing the information about a phenomenon[10]. Using this method, we sought the facts and chronology of events without interpretation. Systematically, we mapped a timeline with each subsequent interview of the interventions and initiatives. Several interviews were necessary in an iterative fashion to clarify details about timelines and interventions. For data triangulation, we reviewed the grey and scientific literature, to include any documented information relevant to the topic.

### Outcome: Provincial 30-day Unplanned All-cause Hospital Readmission Rates

We correlated the initiative timing with the provincial readmission rates using the Discharge Abstract Database (DAD). The DAD records the hospital admission date, the discharge date, the most responsible diagnosis (specified by the hospital attending physician), up to 25 other diagnoses and the acuity (classified as elective or urgent/emergent) of all acute-care admissions to hospitals in the province. This study identified adults aged 20 years or more in the province who were discharged alive after the patient’s first acute-care hospital admission between Jan. 1, 2008, to Dec. 31, 2021, with a primary diagnosis of HF (International Classification of Diseases 10th revision (ICD-10) code I50.x)[11], referred to as the index hospital stay.

The 30-day unplanned all-cause readmission rate represents the proportion of occurrences of a non-elective (unplanned) admission to the hospital for any cause within 30 days of a patient being discharged from the HF index hospital stay. Only the first rehospitalization within 30 days was considered for patients with multiple rehospitalizations. Transfers, sign-outs and deaths within hospitals were excluded. The readmission percentage was calculated as the number of discharges with unplanned readmission to any acute care hospital in Alberta within 30 days from an initial index hospital discharge out of the total number of hospital discharges within the reporting period. Data cannot be shared publicly because to respect the privacy of the participants. Data are available from the University of Calgary Institutional Data Access / Ethics Committee (contact via) for researchers who meet the criteria for access to confidential data.

## Results

### Literature Review Results

Two papers specific to Alberta initiatives were retrieved from the academic literature, including one by Ezekowitz et al.[12], detailing the objectives of the Alberta HEART program, which centred on HF with preserved ejection fraction (HF-PEF). The program sought to develop novel diagnostic criteria and therapeutic approaches for HF-PEF using a multi-phase methodology, that integrates clinical, imaging, and biomarker data. Emphasizing interdisciplinary collaboration and training initiatives, the program aimed to advance translational research in HF. However, while meeting the literature review’s inclusion criteria, the article did not provide results and thus was not included as an initiative within this paper.

The second paper discusses the Alberta Cardiac Access (ACA) initiative, launched in the spring of 2008 in Alberta, aiming to enhance access to specialized HF clinics post-hospital discharge[13]. The ACA initiative was also a topic of discussion in qualitative interviews, elaborated below.

In parallel with database searches, seven web pages were identified and reviewed for relevant content. These web resources were utilized alongside interview responses where applicable. Throughout the paper, relevant content from these documents is incorporated and cited as references.

### Qualitative Results

All informants (n=11) were part of the AHS provincial Cardiac Strategic Clinical Networkᵀᴹ (SCN). Representation from zones was targeted and represented in the sample. Participants included three cardiologists, one general internist, and program directors or project leads employed by AHS during the time of their respective interventions. Given the small sample size, participant details are not provided.

Key informants described five major multi-city/multi-stakeholder initiatives undertaken in Alberta over the years to improve HF outcomes in the province: 1) Alberta Cardiac Access Collaborative initiative which predates the creation of AHS; 2) HF Optimization Project; 3) Provincial Paper Order Set; 4) Electronic Order Set, and 5) Bundles and End-to-End Pathway. All 5 initiatives have used the Discharge Abstract Database (DAD) to measure re-admission rates. The DAD contains demographic, administrative, and clinical data from all Canadian acute care facilities.

#### 1. Alberta Cardiac Access Collaborative (ACAC) initiative

In 2006, the federal government recognized the need for improved access to healthcare and funded several initiatives across the country. The Alberta Cardiac Access Collaborative (ACAC), a multidisciplinary and province-wide group, was formed to improve access to adult cardiac services[7,13]. Between 2006 and 2009, the ACAC created province-wide care delivery models. These models were built upon existing practices used in HF clinics at major city health centers, including Lethbridge, Calgary, Red Deer, and Edmonton. The informant described that “These models informed the creation and expansion of new rural and suburban HF clinics throughout the province and enhanced collaboration with other chronic disease management programs”. All ACAC projects included a multi-phased educational approach, designed to be scaled and spread to other sites. The ACAC projects included HF education materials informed by end-users; a training program for nurses, allied health professionals, physicians, and patients to enhance HF management; improved support for clinical staff at the new clinics (e.g., small-group educational sessions); and opportunities to shadow at the existing clinics. As per the published literature, modest improvements in heart failure readmission rates were observed in ACAC sites, though the extent to which these improvements can be directly attributed to the initiative remains unknown[7,13]. Specifically, after the roll-out of the ACAC initiative, patients discharged from ACAC sites (i.e., those that had specialized HF clinics) exhibited a decrease of 3.6% in 30-day post-discharge death/readmission rates than those discharged from other sites within the province (18.6% versus 22.2%, adjusted odds ratio 0.83, 95% confidence interval, 0.75–0.93). In the South Zone the ACAC, in partnership with the zone leadership, facilitated an integrated care project called the Heart Failure Network (HFN). This multidisciplinary network of healthcare providers followed the integrated Chronic Care Model[14] for services across the continuum of care. The ACAC initiative was the first initiative reported by our key informants, and its associated readmission rates are visible in Figure 3. Some of the gains appeared to be lost over time. According to the key informant on this initiative, the enhanced patient education approach received positive feedback from staff and patients and was integrated into ongoing operational workflows.

**Figure 3.**
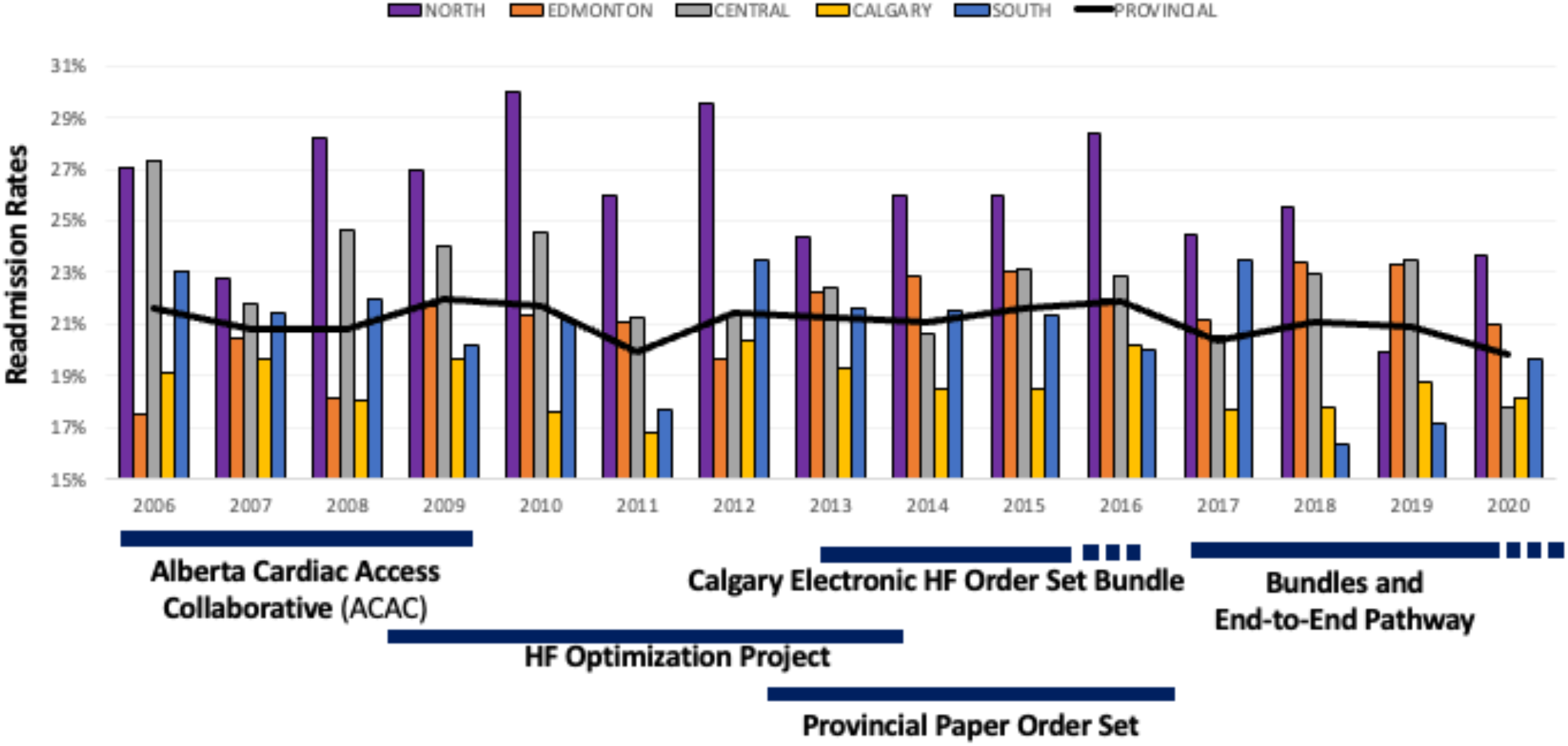
30-Day All-Cause Readmissions for Heart Failure Patients in Alberta by Province and Zones

#### 2. Heart Failure Optimization Project

Building upon ACAC, HF Working Groups were formed across Alberta under the Cardiac Clinical Network to enhance efforts to improve HF care delivery. The groups undertook priority projects, notably the provincial and Calgary Heart Failure Optimization Projects (HFOP).

In early 2008, the provincial HFOP was developed and piloted at Chinook Regional Hospital in Lethbridge. This project operationalized 12 evidence-based best-practice interventions for optimizing inpatient and transitional care for HF patients[15]. By June 2012, seven subsequent facilities across Alberta had implemented one or more interventions based on their resources and patients’ needs (Table 1). Sites that implemented the admission order set, an HF liaison nurse (responsible for facilitating continuity of care for HF patients), and a 3-day follow-up call had the best patient outcomes when assessing length of stay and 30-day all-cause hospital readmission[15]. Although the pilot project demonstrated a reduction in patients’ average length of stay and 30-day readmission rates, 7-day all-cause readmission rates were slightly higher at the end of the evaluation period across all eight sites, highlighting the complexity of achieving sustained improvement across multiple metrics. Again, our trend line shows an initial reduction in readmission followed by a levelling off in average provincial rates (Figure 3).

**Table 1:** Heart Failure Optimization Interventions

|  | Chinook<br>Regional<br>Hospital | Foothills<br>Medical<br>Centre | St.<br>Mary's<br>Hospital | Peter<br>Lougheed<br>Centre | Red Deer<br>Regional<br>Hospital | Rockyview<br>General<br>Hospital | Royal<br>Alexandra<br>Hospital | Mazankowski<br>Heart<br>Institute |
| --- | --- | --- | --- | --- | --- | --- | --- | --- |
| City | Lethbridge | Calgary | Camrose | Calgary | Red Deer | Calgary | Edmonton | Edmonton |
| Project start<br>date | March 1,<br>2008 | October 1,<br>2009 | September<br>1, 2010 | February 1,<br>2011 | June 1, 2011 | March 1,<br>2012 | March 1,<br>2012 | August 1,<br>2012 |
| Notify<br>primary care<br>provider of<br>admission | ✓ | ✓ | ✓ | ✓ | ✓ | ✓ | X | X |
| Provide<br>workbook | ✓ | ✓ | ✓ | ✓ | ✓ | ✓ | ✓ | ✓ |
| Patient<br>education | ✓ | ✓ | ✓ | ✓ | ✓ | ✓ | ✓ | ✓ |
| Notify<br>primary care<br>provider of<br>discharge | ✓ | ✓ | ✓ | ✓ | ✓ | ✓ | ✓ | ✓ |
| Daily weights | ✓ | ✓ | ✓ | ✓ | ✓ | ✓ | ✓ | ✓ |
| Results &<br>discharge<br>summary to<br>patient and/or<br>primary care<br>provider | ✓ | ✓ | ✓ | ✓ | X | ✓ | ✓ | ✓ |
| **Follow-up within 3 days of discharge** | 7-14 days | ✓ | ✓ | X | X | X | X | ✓ |
| **Standard referral process to Cardiac/Heart Function Clinic** | ✓ | ✓ | ✓ | ✓ | ✓ | ✓ | X | X |
| **Admission order set** | X | ✓ | ✓ | ✓ | X | ✓ | X | X |
| **Heart failure liaison nurse receives consults** | ✓ | ✓ | ✓ | X | X | X | X | ✓ |
| **Nurses initiate requests for consults** | ✓ | ✓ | ✓ | ✓ | ✓ | ✓ | ✓ | ✓ |
| **Remind patients to make an appointment with primary care provider before discharge** | ✓ | ✓ | ✓ | ✓ | ✓ | ✓ | ✓ | ✓ |

PCP: Primary Care Provider

#### 3. Provincial Paper Order Set

In 2012, the Cardiac Strategic Clinical Networkᵀᴹ (SCN) developed an HF Order Set to reduce readmission rates for HF patients. SCNs are provincial entities within AHS that strive to provide sustainable access to high-quality care extending from the frontlines to management[16]. SCN actions involve collaboration with zone leadership, operational leaders, clinicians, researchers, and patients across the province on zone-specific priority projects. An order set is a group of standard clinical actions to provide direction to the healthcare team. Order sets are condition-specific and are typically developed from clinical practice guidelines shown to improve patient outcomes[17,18]. The HF Order Set was based on best evidence including the Canadian Cardiovascular Society (CCS) guidelines[19]. This paper-based order set contained all critical elements for evidence-based treatment during an inpatient visit while considering the local resources available.

#### 4. Calgary Electronic Heart Failure Order Set Bundle

The Calgary Zone was charged with piloting the electronic HF order set in Sunrise Clinical Manager™(SCM) the electronic medical record (EMR) system used in three acute care hospitals between 2013 and 2015[20]. With the CCS’ recommendation to leverage the use of technology to improve HF outcomes, we examined how the Calgary zone’s EMR system impacted readmission rates throughout the past 15 years. Calgary Zone’s EMR system, SCM, was used to enhance data-driven projects to tackle HF readmission rates. Predictive analytics were used to monitor outcomes for clinical processes and capture patient feedback. Using machine learning techniques, such as random forest[21,22], the team developed a 30-day readmission prediction model (C-statistic 0.65) coupled with an HF risk stratification tool and embedded it in a web-based dashboard[23]. For each patient admitted, the tool displayed the probability of being readmitted within 30 days of discharge. However, challenges related to the limited integration of the dashboard into clinical workflows, along with average predictive performance, hindered the deployment of these tools to other sites and contributed to a halt in algorithm uptake.

In late 2017, the Calgary HF team made additional efforts to standardize care across all aspects of the HF patient’s journey (from diagnosis to end-of-life) using defined clinical pathways. These included 1) standardizing care in acute care sites, emergency departments, primary care, and multiple community-based services (e.g., outpatient clinics, homecare), and 2) data analytics dashboard for tracking processes and outcome measures. These initiatives are still ongoing, and thus there is limited data available on their effectiveness.

#### 5. Provincial Heart Failure Order Set and Pathway Bundles

Building on the HF Order Set, in 2014 the provincial executive leadership team (AHS CEO and VP) initiated the development of the Provincial HF Pathway. The project was supported by the Cardiac and Respiratory SCNs, and the COPD and HF provincial working groups. It was intended to facilitate the implementation of evidence-based HF care from hospital admission through to discharge and transition into community and primary care (Table 1). Deployment began in 2017, with Red Deer Regional Health Centre as the proof-of-concept site, and 11 other sites following suit. Hence, Central Zone was the first to implement the bundle. The sites could either implement the physician orders and transition to community care orders (Full Bundle) or only implement the transition to community care orders (Transition Bundle) (Table 2). The South Zone implemented the Full Bundle in 2018, followed by Edmonton and North Zones in 2019.

**Table 2:** Components of the Heart Failure Order Set Bundle

| Components | Description | Completed by |
| --- | --- | --- |
| Physician Admission Orders | Evidence based acute admission order set recommendations | Physician |
| Transition to Community Care | Recommended consultations, patient education, discharge plan, and follow-up | Pre- Populated |
| LACE Index Scoring Worksheet | Tool to identify risk of readmission to hospital within 30 days of discharge | Health Care Provider(s) |
| Risk Stratification Worksheet | Algorithm to identify the recommended time until follow-up with Heart Function Specialist/Clinic and with Primary Care Clinic based on patient risk. | Health Care Provider(s) |
| Admission to Discharge Checklist | Tool to assist staff identify and record completion of activities related to HF patient care | Health Care Provider(s) |
| Discharge Management Plan | Resource to review with HF patient prior to hospital discharge. Identifies key messages, resources and follow up information. Personal assessment to be completed by patient.<br>Provide copy to patient upon discharge | Health Care Provider and Patient |

HF: Heart Failure; LACE: Length of stay, Acute/emergent admission, Charlson comorbidity index score, Emergency department visits in previous 6 months

In 2019, the provincial HF Working Group became the HF Care Path team and updated the paper order set. The team also updated the electronic version of an HF Care Path while AHS began deployment of a large-scale EMR integration project, Connect Care, to streamline patient care, through the sharing of health information. This unified clinical information system, Epic^TM^, will replace the 1,300 systems across AHS and affiliated healthcare organizations. The project will also better align paper-based clinical pathways and order sets with electronic workflows, thus allowing for more standardized care and potentially improved outcomes.

## End-to-End (E2E) Pathway

In 2018, with the achievements and experiences acquired through the projects, AHS, in collaboration with the Cardiovascular Health and Stroke SCN ™, launched a 3-year plan for an End-2-End Clinical Care Pathway for HF. This pathway integrates the entire care continuum in Alberta. The pathway includes the following elements in hopes of improving patient outcomes: 1) a mapping of the HF patient journey within acute care, including patient stories as they transition from home to hospital and back to home, and 2) an in-depth analysis of the factors underpinning predictive modelling of patient prognosis and death. Outcomes need yet to be measured.

In 2019, the provincial HF Working Group became the HF Care Path team and updated the paper order set. The team also updated the electronic version of an HF Care Path while AHS began deployment of a large-scale EMR integration project, Connect Care, to streamline patient care, through the sharing of health information. This unified clinical information system, Epic^TM^, will replace the 1,300 systems across AHS and affiliated healthcare organizations. The project will also better align paper-based clinical pathways and order sets with electronic workflows, thus allowing for more standardized care and potentially improved outcomes.

In summary, Figure 3 demonstrates that there is no consistent trend within zones according to the interventions implemented in Alberta. Modest decreases in provincial readmission rates were observed with each new intervention, suggesting potential benefit. Additionally, over the 14 years depicted in Figure 3, a 1.8% reduction in provincial readmission rates was observed, particularly with more comprehensive initiatives like the End-to-End pathway. However, the specific contribution of these initiatives to the overall trend remains difficult to determine given the multifactorial nature of healthcare delivery.

## Discussion

The multifactorial nature of readmissions highlights involved in effectively addressing the issue. Alberta Health Services has implemented several interventions aimed at improving patient care and outcomes in HF. These interventions have relied heavily on strong leadership, clinician engagement, and integration of electronic-based tools. However, challenges such as fragmented resource allocation, variability in intervention implementation, and suboptimal communication between healthcare teams may undermine the potential impacts of the interventions on reducing HF readmission rates. The transition from paper-based to electronic order sets in Calgary represents a step towards addressing these challenges, with some lower readmission rates seen.

The impact of interventions varied across different zones within Alberta, likely influenced by demographic disparities. Urban centers like Calgary and Edmonton, characterized by commuter communities, exhibited outcomes that differed from those in rural areas, which often face limited healthcare resources (e.g., fewer HF clinics and cardiologists). Despite these variations, combined interventions seemed to be associated with some reductions in readmission rates. While a previous clinical trial[24] has suggested minimal reductions in HF readmission rates, the iterative, multi-phased, and multi-sectorial nature of the interventions discussed here, along with the implementation of an integrated EMR system to streamline province-wide HF care, presents promising opportunities for improving HF outcomes on a broader level.

## Personalized HF Care – the Future in Alberta

The future of HF care in Alberta is marked by a shift towards personalized approaches facilitated by the Connect Care clinical information system. Predictive analytics will enable the identification of high-risk patients, guiding tailored post-hospital outpatient follow-up. As Connect Care expands, evidence-based technology-driven initiatives, such as HF dashboards, order sets, and pathways will be scaled to spread successful local initiatives to the whole province. Alberta has the advantage of both central and local clinical teams, partnered with operational and academic leaders to keep the quality of HF care moving forward. In addition, other initiatives include a personalized medication titration framework using cluster scheme approaches to enhance patient outcomes[25]. Both data-driven and clinically personalized treatments offer future promise for patients with HF.

## Strengths and Limitations

The information gathered in this environmental scan incorporated verbal and written accounts from multiple stakeholders related to HF clinical care and research. While snowball sampling facilitated the identification of key informants, it may have limited the diversity of perspectives presented. However, all 11 participants were selected due to their significant involvement from the beginning and their participation in most, if not all, interventions, ensuring comprehensive insights.

The reliance on historical recall was mitigated by confirming information from multiple sources. While some outcome data was available, a lack of detailed data related to specific interventions limited the ability of teams to evaluate their efforts and limited what could be reported. Further, changes in medical therapies or vascular procedures occur simultaneously with non-invasive interventions. This poses a challenge in assessing the isolated impact of these interventions. Therefore, the qualitative nature of an environmental scan does not allow for quantitative analysis of the potential confounding effects of medical advancements in HF management over the study period. Similarly, advances in medical care for HF, while primarily for chronic HF, could be contributing to the change in readmission rates over the study period. Going forward, coordinated provincial electronic data dashboards will enhance monitoring interventions and outcomes for HF care. Lastly, the effect of interventions on HF readmissions could be better estimated with the use of interprovincial data comparisons. Though an attempt was made to request comparative data from British Columbia and Ontario, the attempt was unsuccessful. Further efforts will be made to encourage data sharing to compare provincial HF readmission data and initiatives.

## Relevance to Stakeholders

The findings from this environmental scan carry significant implications for various stakeholders. For clinicians, the integration of electronic tools and standardized order sets can streamline workflows, improve communication, and enhance patient outcomes. Policymakers may utilize these insights to inform provincial and national policies aimed at improving HF care quality and reducing hospital readmissions, while also considering funding models that support interdisciplinary care delivery. For health system administrators, the study highlights the importance of optimizing resource allocation and fostering collaboration between urban and rural settings, alongside the need for ongoing quality improvement initiatives and the establishment of performance metrics to track progress over time.

## Conclusions

In summary, this environmental scan identifies various comprehensive initiatives in Alberta aimed at enhancing patient care and reducing HF readmission. Over the past 15 years, a 1.8% reduction in 30-day all-cause readmission rates was observed; however, attributing this to improvement directly to the interventions remained challenging given the complex nature of healthcare delivery and multiple influencing factors. Continued efforts focused on personalized care and the optimized use of EMR systems hold promise for further improvement in HF readmission rates, although ongoing monitoring and assessment will be critical to fully understand their impact.

## Data Availability

Data cannot be shared publicly because to respect the privacy of the participants. Data are available from the University of Calgary Institutional Data Access / Ethics Committee (contact via) for researchers who meet the criteria for access to confidential data.

https://www.albertahealthservices.ca/assets/zone/ahs-map-ahs-zones.pdf

## Acknowledgments

The authors are grateful to the members of the Cardiac Strategic Clinical Network, the Heart Function Clinic AHLP Southwest Zone team, and Alberta Health Services for their support and help in gathering the information necessary for the completion of this article.

